# Trends in NIH funding for pediatric research, FY2020–FY2026

**DOI:** 10.64898/2026.06.29.26356863

**Authors:** Fred D. Ledley, Reagan Mozer

**Affiliations:** Center for Integration of Science and Industry, Bentley University, Waltham, Massachusetts; Departments of Natural & Applied Sciences, Department of Management, Bentley University, Waltham, Massachusetts; Department of Mathematical Science, Bentley University, Waltham, Massachusetts

## Abstract

There were substantial changes in NIH policies regarding research funding in FY2025. This work examines NIH funding for pediatric research FY2020 through Q2FY2026 including the number and cost of awards, the number of first year (type 1) awards, the number of Notices of Funding Opportunity, and the topical focus of research awards. NIH funding for pediatric research declined >20% in the first two quarters FY2025-FY2026 with proportionally greater reductions in first year awards and Notices of Funding Opportunity. Changes were also noted in the topic prevalence of new awards consistent with 2025 guidelines identifying topics “not aligned with NIH priorities.” These results suggest that pediatric research aimed at advancing healthcare for children is at risk with potential collateral consequences beyond childhood.

## Introduction

Following changes in NIH policies starting in the second quarter of FY2025, the number of research project awards declined by 20.5% in FY2025^1^ with further reductions in the first half of FY2026. ^2^ This work examined pediatric research awards over this period in the context of longstanding concerns regarding the adequacy of NIH support for pediatric research. ^3–6^ Specifically, we assessed trends in (1) the number and cost of awards involving pediatric research, (2) the number of first year (type 1) awards; (3) the number of Notices of Funding Opportunity for pediatric research, and (4) the topical focus of awards in the first half of fiscal years FY2020 through FY2026.

## Methods

### Study Design

Cross-sectional study of NIH funded projects awarded from FY2020 through Q2FY2026.

### Data

Project awards and costs were identified in NIH RePORTER (accessed 4/23/2026) with the search “pediatric OR child OR childhood OR infant OR newborn” limited to Project Title, Project Terms, and Project Abstract fields. Fiscal quarters were assigned using the “Award Notice” date. Notices of Funding Opportunity for pediatric research were identified in grants.gov or NIH Guide using the same Boolean search strategy. Fiscal year quarters were assigned using the “Posting” or “Release” Date.

### Analysis

To evaluate changes in topical focus among newly funded projects, we analyzed Public Health Relevance narratives from type 1 (“new project”) awards. For each fiscal year, duplicate and near-duplicate narratives were collapsed to retain one narrative per award, yielding a corpus of 14,788 unique narratives. We fit a 30-topic structural topic model (STM)^7^ with fiscal year included as a document-level covariate allowing topic prevalence to vary by fiscal year and estimated the proportion of each narrative assigned to each topic. Topics are summarized with frequency-exclusivity (FREX) terms^8^, which identify words that are common within and relatively exclusive to each topic. The number of topics (K=30) was selected from a diagnostic sweep across a grid of candidate values based on held-out likelihood, semantic coherence, exclusivity, and residual dispersion. Additional details are provided in Supplemental Material (attached).

### Limitations

This analysis was not based on the Research, Condition, and Disease Categorization (RCDC) system commonly used to identify classes of research funding. A recent NAS report challenged the utility of RCDC for identifying pediatric research,^5^ pointing out specific deficiencies as well as the lack of transparency and documentation. The dataset identified by Boolean search, while transparent, is used as an indicator of pediatric research support and does not include all NIH funded research with pediatric applications.^5^

## Results

### Project awards and costs

The study dataset included 66,739 NIH project awards totaling $47.7 billion from FY2020 through Q2FY2026 with 20,468 awards totaling $12.8 billion in the first two quarters of these fiscal years. During the first two quarters of FY2020 through FY2024, the average number of awards was 3,197 with average NIH costs of $1.97 billion.

During the first two quarters of FY2025 there was an average of 2,515 awards with average NIH cost of $1.6 billion. This represents a decline of 21.3% in the number of awards and 18.0% in NIH costs, compared to the FY2020-–FY2024 average. During the first two quarters of FY2026, there were 1,968 awards with NIH costs of $1.28 billion. This represents a decline of 38.4% in award number and 35.3% in NIH costs relative to FY2020–FY2024 (Figure 1A-B).

**Figure 1.**
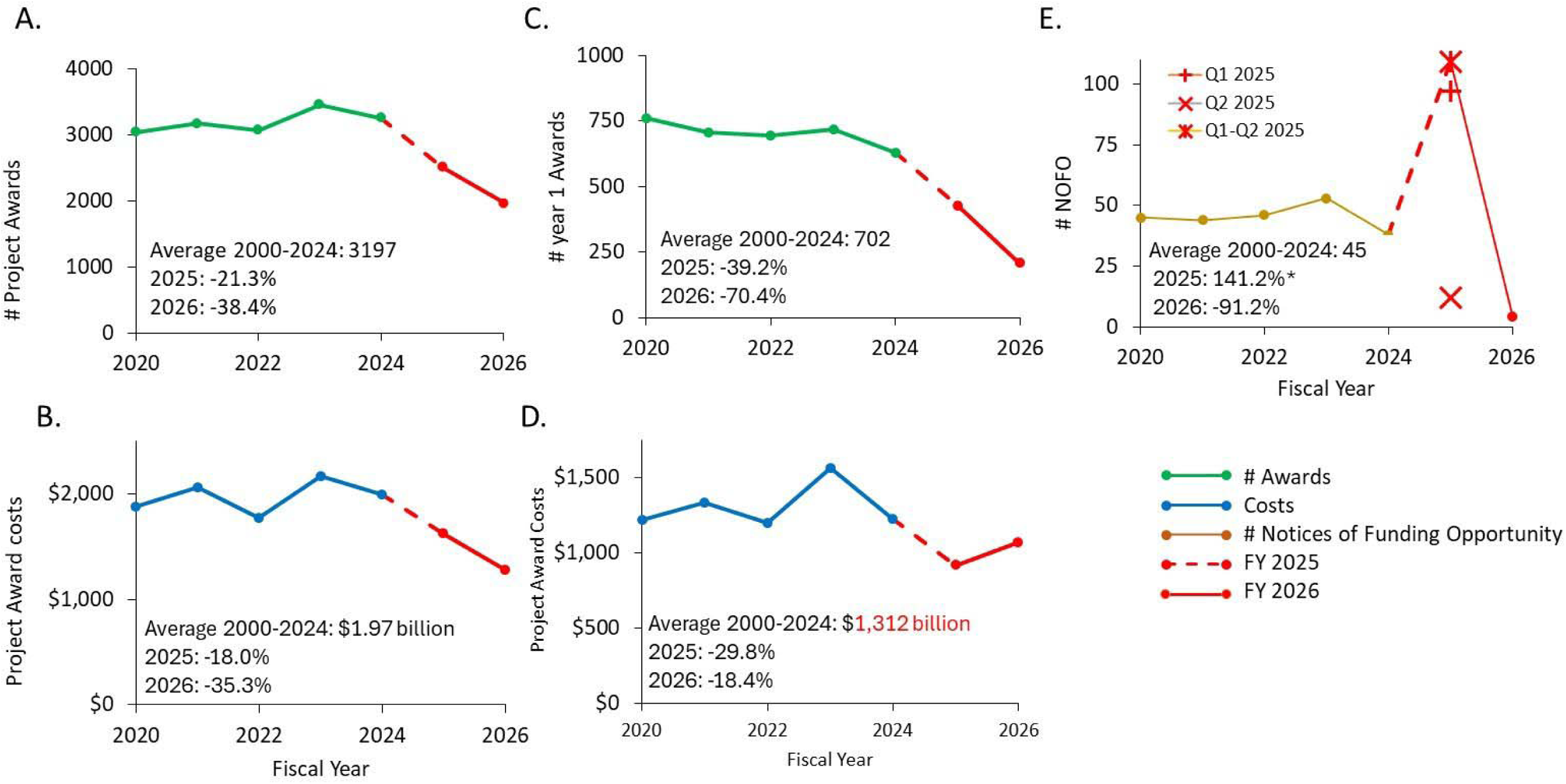
NIH Project Awards and number of Notices of Funding Opportunity involving pediatric research in the first two quarters of FY2020–FY2026. A. Number of project awards; B. Award costs. C. Number of first year Project Awards (new projects); D. Costs associated with first year Project Awards; E. Number of Notices of Funding Opportunity. Points represent annual values. The average from FY2020 to FY2024 and the % reduction in f FY2025 and FY2026 are shown in each panel. *The increased number of Notices of Funding Opportunity in the first two quarters of FY2025 included a record high number of Notices in Q1 2025 and then-record low number of Notices in Q2 2025. Award costs are reported in millions of US dollars and were not adjusted for inflation.

During the first two quarters of FY2020 through FY2024, the average number of first year awards was 702 and the average NIH cost was $1.31 billion. In the first two quarters of FY2025, there were 416 first year awards with an NIH cost of $0.92 billion. This represents a decline of 39.2% in award number and 29.8% in award costs. There were 208 first year awards in Q1-Q2 FY2026 with NIH costs totaling $1.0 billion), representing 70.4% in award number and 18.4% in award costs. (Figure 2C-D)

**Figure 2.**
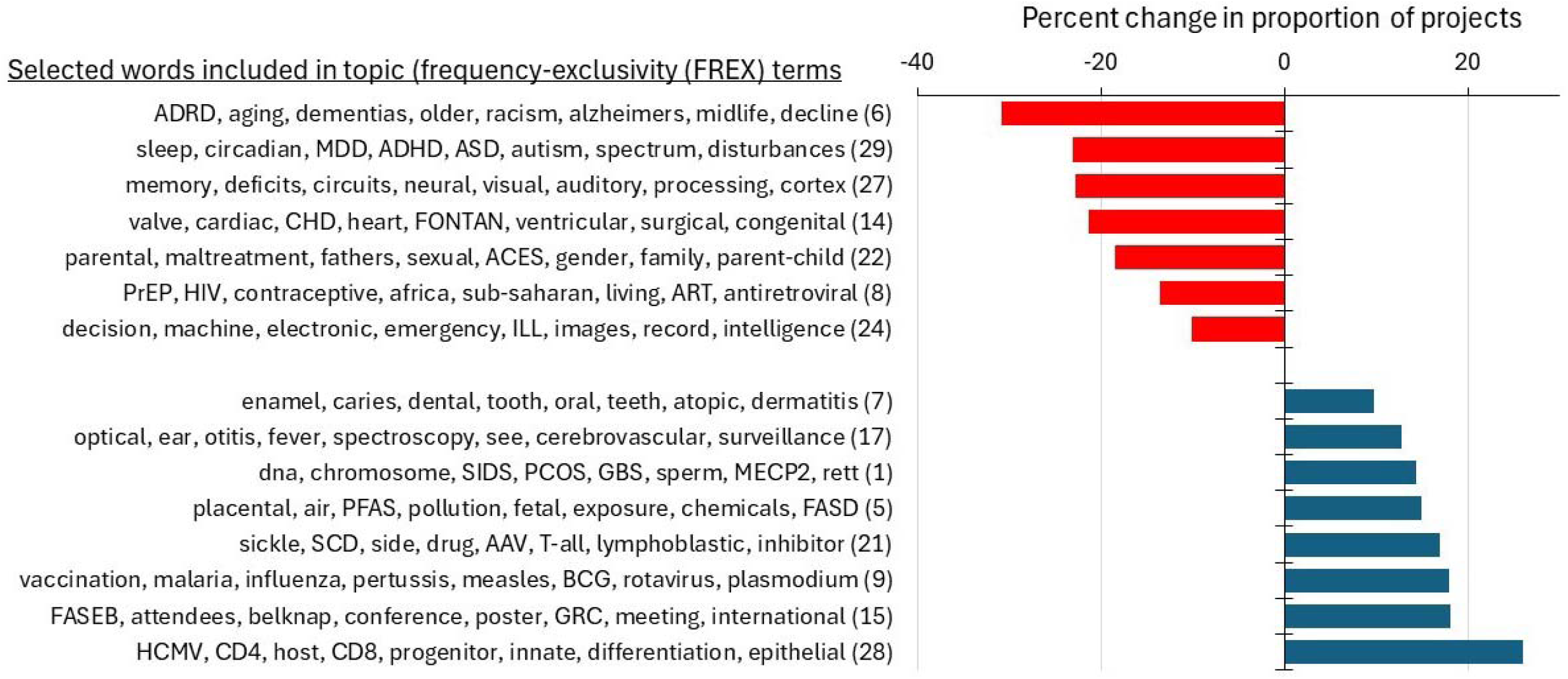
Relative changes in topic prevalence FY2025–FY2026 compared to FY2020–FY2024 baseline. Bars show relative percent changes in mean document-level topic proportions for topics with changes >10% relative to the FY2020-FY2024 baseline. Labels show the top 8 de-duplicated FREX terms^8^ for each topic, representing words that are statistically common within, and exclusive to, that topic. The full table of all 30 topics is provided in the Supplemental Material.

### Notices of Funding Opportunity

There was an average of 45 Notices of Funding Opportunity for pediatric research in the first two quarters from FY2020 through FY2024. There were 109 Notices of Funding Opportunity in the first two quarters of FY2025, including 97 notices in Q1 (September-December 2024) and 12 in Q2 (January-March 2025). There were only 4 pediatric-related notices in the first two quarters of FY2026, a 91% decrease relative to the FY2020–FY2024 baseline. (Figure 1E)

### Topic prevalence in awards FY2020–FY2024 and FY2025–FY2026

The document-level topic database is described in Supplemental Material (Table S1, attached). Mean document-level topic prevalence in FY2025–FY2026 differed from the FY2020–FY2024 baseline. Fifteen of the thirty identified topics demonstrated relative changes exceeding 10% in mean document-level prevalence (Figure 2). Several declining topics include FREX terms that also appeared in the 2025 NIH guidance on awards “…not aligned with the NIH’s priorities.”^10^ For example, “racism” appeared among FREX terms within a topic also characterized by the terms “Alzheimer’s” and “aging.” Similarly, “gender” appeared within a topic also characterized by “fathers” and “parent-child.” This analysis shows that words subject to the 2025 NIH guidelines as being inconsistent with “NIH priorities” occurred with within broader scientific topics that included words not governed by this guidance. This suggests that administrative screening based on individual terms could affect multiple projects spanning multiple areas of research. Additional data on the FREX terms associated with each topic and topic prevalence estimates are provided in the Supplemental Material (Table S2, attached).

## Discussion

These data identified substantive declines pediatric research awards, first-year awards, and pediatric-related Notice of Funding Opportunity in the first two quarters of FY2025 and FY2026 compared to the FY2020 through FY-2024. The greatest proportional reductions were observed for first-year awards and Notices of Funding Opportunity.

The disproportionately larger decline in first-year awards relative to the total number of awards is notable because these awards represent the entry point for multi-year research programs. Reductions in first-year funding may have effects extending beyond the fiscal years examined here by reducing the pool of projects eligible for continuation and renewal in future years. Moreover, the marked reduction in pediatric-related Notices of Funding Opportunity in the first two quarters of FY2026 is indicative of a breakdown in the process by which the NIH has historically identified priority areas for pediatric research and solicited investigator-initiated projects in those areas. These findings raise concern about the future of NIH funding related to pediatrics, children, childhood, infants, and newborns and suggest the future of federal funding for pediatric research is at risk.

The observed changes in the topic prevalence of newly funded awards are consistent with the 2025 NIH guidelines, which identified selected topics as “not aligned with NIH priorities” and reduced funding for those topics. Thus, the observed changes in topic prevalence are best interpreted as evidence of a shift in the focus of pediatric research funding rather than the scientific merit of individual research awards. These changes are of particular concern not only because certain topics that are important in pediatrics may be explicitly censored, but also because of the potential for collateral effects that decrease NIH funding for research in areas described by statistically associated terms.

These results should be interpreted in the context of broad changes in NIH research funding since the second quarter of FY2025.^1,2^ The observed decreases in awards for pediatric research and Notices of Funding Opportunity are consistent with broader trends in NIH research funding since that time.^1,2,9^ (See tracking by AAMC: https://www.aamc.org/about-us/biomedical-research/publication/tracking-nih-funding). Further studies are required to assess whether changes in NIH support for pediatric research are comparable to changes affecting other areas of research. In light of longstanding concern about the adequacy of NIH support for pediatric research,^3–5^ continued surveillance is warranted to assess the impacts of continuing change in NIH policy on pediatric research that necessary to advance health care for children and its consequences beyond childhood.

## Data Availability

All data in the present work are available from public databases without restriction.

## Author contributions

*Study concept and design:* Ledley

*Acquisition, analysis, and interpretation of data:* Ledley, Mozer

*Drafting of the manuscript:* Ledley, Mozer

*Critical revision of the manuscript for important intellectual content:* Ledley, Mozer

*Obtained funding:* Ledley

*Administrative, technical, or material support:* Ledley

*Supervision*: Ledley

## Author Access to Data

Ledley had full access to all of the data in the study and takes responsibility for the integrity of the data and the accuracy of the data analysis.

## Conflict of Interest Disclosures

All authors have submitted ICMJE Forms for Disclosure of Potential Conflicts of Interest.

## Funding/Support

This work was supported by a grant from National Biomedical Research Foundation to Bentley University.

## Role of the Sponsor

The sponsors had no role in the design and conduct of the study; collection, management, analysis, and interpretation of the data; preparation, review, or approval of the manuscript; or decision to submit the manuscript for publication. The sponsors did not have the right to veto publication or to control the decision regarding which journal the paper was submitted.

## Non-author Contributions to Data Collection, Analysis, or Writing/Editing Assistance

n/a

## Data Sharing Statement

All data used in this study are publicly available without restriction from the following sources: NIH RePORTER: https://reporter.nih.gov/; Grant.gov: https://www.grants.gov/search-results-detail/349644; NIH.gov https://grants.nih.gov/funding/nih-guide-for-grants-and-contracts

## Supplemental Material - Topic prevalence

### Data Cleaning/Preparation Procedure

1. Remove boilerplate labels. Convert blank, n/a, na, none, null, and 0 narrative values to missing.
2. Clean the narrative text by removing line breaks, squishing repeated whitespace, converting non-ASCII characters to ASCII where possible, and removing other nonstandard/unrecognizable characters.
3. Keep rows with nonmissing raw relevance text and nonmissing type variable

- Exclude any with <20 words after cleaning
- Exclude rows with average word length > 10 characters (these had conversion issues and the text is not legible)
4. Screen for identical or near-duplicate cleaned narratives and flag rows where text is duplicated (or very close) in other rows in the data set
5. Create a project/type spine by selecting the earliest available record within each core project number and award type, ordered by fiscal year and award notice date. This gives the first observed record for each project/type combination.
6. Filter to only Type 1 awards.
7. Remove duplicated narratives by keeping the one with the earliest award notice date.
8. Build a type 1 award corpus from the deduplicated Type 1 records. Tokenize the cleaned relevance text by removing punctuation, numbers, and symbols; lowercase terms; remove English stopwords plus project-generic terms and extremely common terms (i.e., terms appearing in >50% of documents)

**Table S1.** Topic Preference Data Description.

| **Table S1. Topic Preference Data Description** |  |
| --- | --- |
| **Metric** | **Count** |
| Raw workbook rows | 83,193 |
| Rows retained after initial data/text cleaning (step #4) | 71,658 |
| Screen for unique relevance text (step 5) | 26,837 unique texts |
| Project/type spine rows (step 6) | 40,817 unique project number-type combinations |
| Filter to Type 1 projects (step 7) | 14,882 |
| Deduplicate Type 1 projects with identical or near-identical text (step 8) | 14,793 |
| Remove documents with no observed text after removing stopwords and extremely common terms | 14,788 |
| **Final Dataset Counts** |  |
| STM input documents | 14,788 unique Type 1 projects with relevance text available |
| STM vocabulary terms | 80,307 unique words used across input documents |

### Text Cleaning & Pre-processing

Text cleaning was performed using R to standardize the structure and formatting of all project narratives while preserving the content of each narrative. This included removing common labels and administrative boilerplate including variations of the terms “Project Narrative,” “Public Health Relevance,” “Relevance,” and “Narrative,” converting line breaks to spaces, stripping repeated whitespace, removing leading punctuation, and converting all text to ASCII encoding using Unicode NFKC normalization. After cleaning, rows were included in the analysis only if the original narrative text (Public Health Relevance) relevance field was nonmissing, the cleaned narrative contained more than 20 words, and average word length was less than 10 characters. This allowed us to remove cases where the narrative text contained only short text snippets (with no meaningful topical information) as well as rows containing corrupted or concatenated text.

Prior to analysis, the cleaned relevance narratives were converted to a document-feature matrix and tokenized to remove punctuation, numbers, and symbols. Tokens were converted to lowercase and screened to remove standard English stopwords, project-generic terms (e.g., “research,” “study,” “project,” “aim”), words shorter than 3 characters, and extremely common terms (i.e., words appearing in more than 50% of documents. The resulting corpus contained 14,788 documents and 80,307 unique vocabulary terms.

### Model Fitting & Analysis

Using the final corpus of cleaned narrative texts, we fit a structural topic model (STM), an extension of Latent Dirichlet Allocation that allows topic prevalence to vary as a function of document-level covariates. The primary model specified K=30 topics and modeled topic prevalence as a function of fiscal year. Model fitting was performed using the stm package in R (Roberts et al., 2019).

From the fitted STM, we computed document-level topic proportions estimating the proportion of content within each document attributable to each of the 30 identified topics. Document-level proportions were then aggregated by topic and fiscal year to compute the average topic share among Type 1 projects funded in each fiscal year. Period averages were then compared for projects funded in fiscal years 2025-2026 with those from 2020-2024, with relative percent changes in each topic calculated as 100 x (recent period mean – baseline period mean) / baseline period mean.

The table below shows the top 8 Frequency-Exclusivity (FREX) terms for each of the 30 identified topics as well as the average document-level topic proportions for Type 1 projects funded in each period and the percent change in topic share between periods.

**Table S2.**
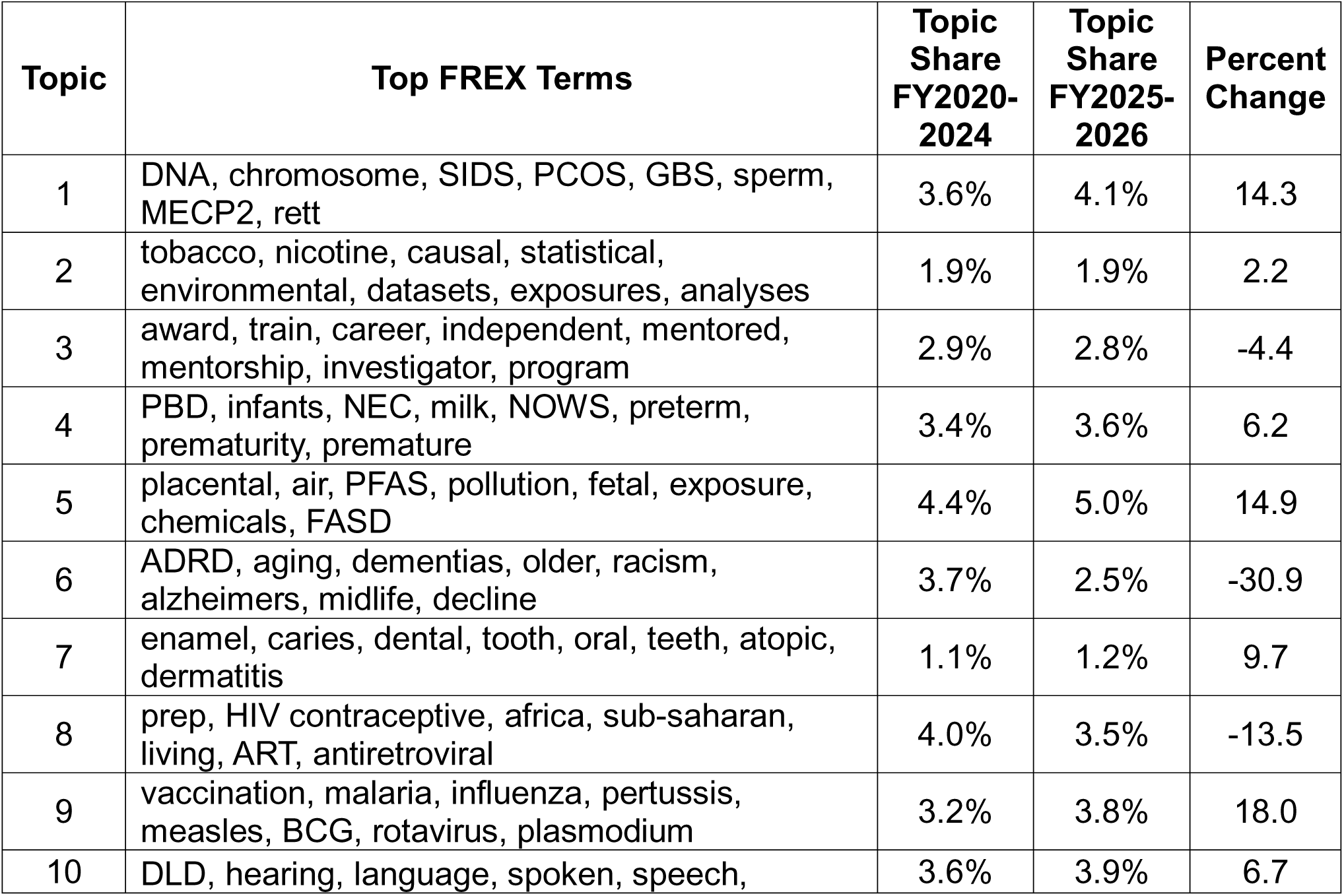
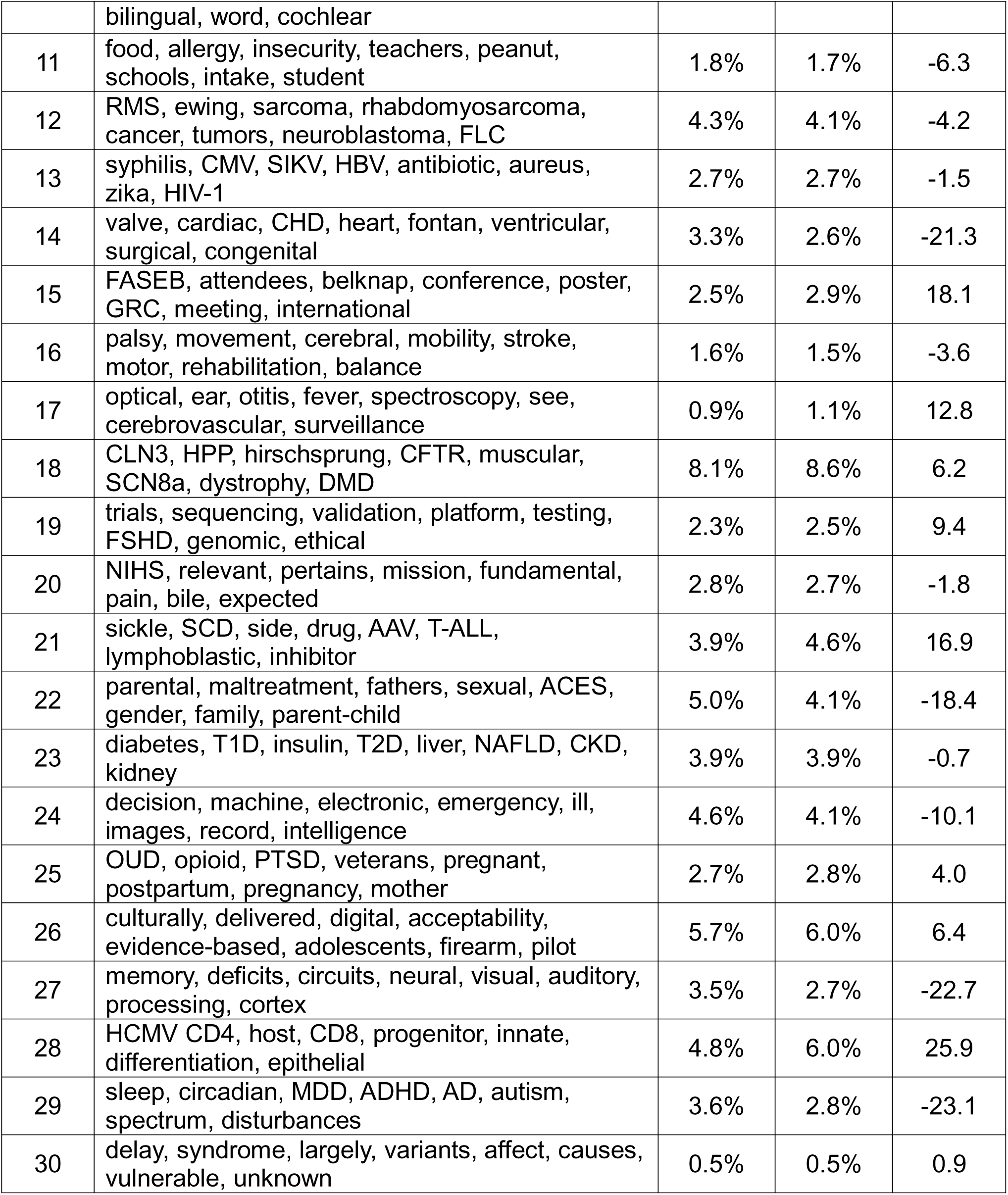
Topic Preference Detailed Results.

## Notes

### Competing Interest Statement

Dr, Mozer has received research funding to Bentley University from the Institute of Education Sciences, US Department of Education and from the VA Health Services Research and Development. Dr. Ledley has received research funding to Bentley University from the National Biomedical Research Foundation (non-profit).

## References

1. NIH. Fiscal Year 2025 by the numbers: extramural grant investments in research. 2026. March 12, 2026. https://grants.nih.gov/news-events/nih-extramural-nexus-news/2026/03/fiscal-year-2025-by-the-numbers-extramural-grant-investments-in-research

2. Mueller B, Hwang I. Pace of NIH funding slows further in Trump’s second year. The agency has approved far fewer new grants than it did in years past. A renewed effort to screen for disfavored terms and a loss of personnel are contributing. NY Times. 5/2/2026. https://www.nytimes.com/2026/04/22/science/trump-nih-funding-research.html

3. Gitterman DP, Hay WW. That sinking feeling, again? The state of National Institutes of Health pediatric research funding, fiscal year 1992–2010. Pediatric research. 2008;64(5):462-469. https://www.nature.com/articles/pr2008226

4. Gitterman DP, Langford WS, Hay Jr WW. The fragile state of the National Institutes of Health pediatric research portfolio, 1992-2015: doing more with less? JAMA pediatrics. 2018;172(3):287-293. https://jamanetwork.com/journals/jamapediatrics/fullarticle/2668294

5. Dennery P, Rivara F, Wojtowicz A, Perera U, National Academies of Sciences E, Medicine. Strategies to Enhance NIH-Funded Pediatric Research: Optimizing Child Health. 2026; https://www.aps1888.org/wp-content/uploads/2026/01/NASEM-NIH-Peds-Research-Full-Report.pdf

6. Congress. Children’s Health Act of 2000, Public Law 106–310. In: Congress U, editor. 2000. https://www.congress.gov/bill/106th-congress/house-bill/4365

7. Roberts ME, Stewart BM, Tingley D, et al. Structural topic models for open-ended survey responses. American journal of political science. 2014;58(4):1064–1082. https://onlinelibrary.wiley.com/doi/abs/10.1111/ajps.12103

8. Bischof J, Airoldi EM. Summarizing topical content with word frequency and exclusivity. 2012:201–208. https://dl.acm.org/doi/abs/10.5555/3042573.3042578

9. E G. The NOFO graveyard; a response to NIH’s defense of its vanishing funding opportunities. 2026. https://elizabethginexi.substack.com/p/the-nofo-graveyard

10. Staff Guidance - Reviewing Grants for Priority Alignment, Issue Date: December 20, 2025 (copy published by AAAS) (AAAS) (2025). https://www.science.org/pb-assets/PDF/News%20PDFs/final_staff_guidance-1765918233.pdf

## Supplemental material – Reference

Roberts, M. E., Stewart, B. M., & Tingley, D. (2019). Stm: An R package for structural topic models. Journal of statistical software, 91, 1–40.

